# Double Invisible Stigma in Young Adults with PFO-Related Stroke: A Qualitative Study

**DOI:** 10.1101/2025.08.18.25333728

**Authors:** Chen Chen, Ting Xu, Zhujun Zeng, Ziyu Liu, Yuankai Li, Jing Miao, Rong Hu, Yiming Gao, Meng Li, Chunlin Li, Xiaoli Yang

**Affiliations:** Department of Cardiac Surgery, Huashan Hospital, Fudan University, Shanghai, China; Department of Hospital Infection Control, Huashan Hospital, Fudan University, Shanghai, China; Department of Gastroenterology, Huashan Hospital, Fudan University, Shanghai, China; Taizhou People’s Hospital Affiliated to Nanjing Medical University, Taizhou, Jiangsu, China; School of Nursing, Fudan University, Shanghai, China

**Keywords:** patent foramen ovale, cryptogenic stroke, stigma, young adults, grounded theory, qualitative research

## Abstract

**BACKGROUND AND PURPOSE:** Patent foramen ovale (PFO) accounts for 40% of cryptogenic strokes in adults under 60 years. While percutaneous closure effectively prevents recurrence, psychosocial impacts remain unexplored. We aimed to develop a theoretical framework for understanding stigma experiences in young adults with PFO-related stroke.

**METHODS:** Using constructivist grounded theory, we interviewed 26 young adults (median age 42.5 years) who underwent PFO closure following cryptogenic stroke at a tertiary center in China (2022-2024). Semi-structured interviews explored stigma experiences. Data were analyzed using constant comparative method until theoretical saturation.

**RESULTS:** Analysis revealed "double invisible stigma", a novel phenomenon where intersection of congenital cardiac defect invisibility and age-inappropriate stroke creates compound psychological distress exceeding either condition alone. Five themes emerged: emotional landscape of compound stigma, social consequences and relational disruption, multilevel stigma sources, coping strategies and adaptation, and healthcare navigation challenges. A temporal evolution model identified three phases: Acute Bewilderment (0-3 months), Strategic Navigation (3-9 months with peak stigma), and Crystallized Duality (9+ months). High-stigma participants showed 2.3-fold higher healthcare utilization and 75% longer work disability duration. We developed the Double Invisible Stigma Screen (DIS-Screen) for clinical assessment.

**CONCLUSIONS:** Young adults with PFO-related stroke experience unique compound stigma with predictable temporal evolution. The 3-9 month window represents critical intervention opportunity. Integration of stigma screening and phase-specific support into PFO-stroke programs may improve psychological and healthcare outcomes.

---

Patent foramen ovale (PFO) accounts for up to 40% of cryptogenic strokes in adults under 60 years.^1-3^ Recent trials establishing percutaneous closure’s superiority over medical therapy have transformed PFO-related stroke into a treatable condition.^4-6^ Yet the lived experiences of these young stroke survivors remain poorly understood.

Young adults with PFO-related stroke face a peculiar paradox. They must process having both a congenital heart defect, discovered only after their stroke, and a neurological event typically associated with aging. This dual diagnosis disrupts core assumptions about health and identity during life stages characterized by career building and family responsibilities.^7, 8^

Stigma profoundly shapes illness experiences. Link and Phelan’s framework^9^ describes how labeling and stereotyping lead to status loss and discrimination. While stroke stigma has been documented,^10-12^ existing work focuses on older adults with visible disabilities. We know little about stigma when both the cardiac anomaly and stroke sequelae remain largely invisible to others.

This invisibility may complicate recovery in unexpected ways. Without visible markers of illness, young PFO-stroke patients often encounter skepticism about their limitations or struggle to access support. Simultaneously, having a stroke decades "too early" violates social expectations about who gets sick and when. These intersecting challenges likely influence treatment adherence, work participation, and healthcare utilization,^13-15^ yet remain unexamined.

We conducted a grounded theory study to understand how young adults experience stigma after PFO-related stroke. Our goal was to develop a theoretical framework that could guide clinical interventions for this growing population.

## METHODS

### Study Design

We used constructivist grounded theory^16^ to explore stigma experiences in young adults with PFO-related stroke. This approach was selected to develop theoretical understanding of how patients navigate the dual diagnosis of congenital heart condition and stroke.

### Sample and Recruitment

The study was conducted within the multidisciplinary PFO-stroke program at Huashan Hospital, Fudan University, Shanghai, China (the Multidisciplinary Care Pathway, see Figure 1). Inclusion criteria were: (1) adults aged 18-55 years diagnosed with cryptogenic stroke and PFO; (2) stroke within the past 5 years; (3) ability to communicate in Mandarin; and (4) no severe cognitive impairment. We used purposive sampling to ensure variation in age, gender and treatment decisions. The hospital’s stroke nurse coordinator identified eligible patients and obtained consent for researcher contact. Recruitment continued until data saturation at 26 participants. ( Study Flow Diagram, see eFigure 1)

**Figure 1.**
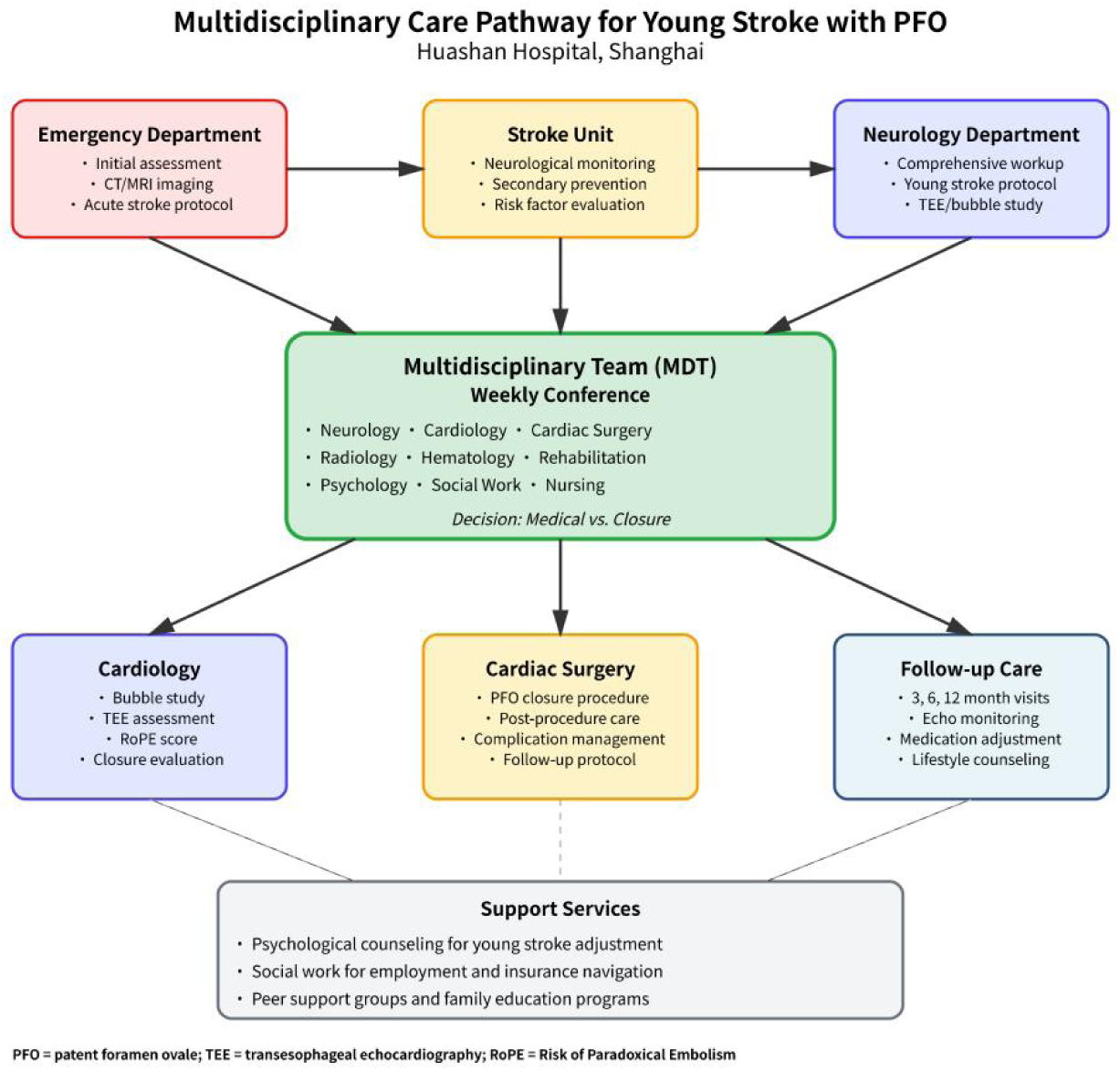
Multidisciplinary Care Pathway for Young Adults with Patent Foramen Oval e-Related Stroke. Flowchart depicting the integrated care pathway at Huashan Hospital, Shanghai. Patients progress from Emergency Department through Stroke Unit to Neurology Department for comprehensive evaluation including transesophageal echocardiography (TEE) and bubble study. Weekly multidisciplinary team conferences involving specialists from 9 disciplines determine treatment decisions (medical management vs. PFO closure). Based on team recommendations, patients receive specialized care through Cardiology (evaluation and medical management), Cardiac Surgery (PFO closure), or structured Follow-up Care (3, 6, and 12-month visits). Support services address psychosocial needs thro ugh counseling, social work assistance, and peer support programs. PFO = patent fora men ovale; RoPE = Risk of Paradoxical Embolism; TEE = transesophageal echocardio graphy.

### Data Collection

Individual semi-structured interviews were conducted by two researchers trained in qualitative methods. Twenty interviews occurred face-to-face in a meeting room, six were conducted via video call for geographically distant participants. Interviews lasted 45-90 minutes and were audio-recorded with written consent. The interview guide (see eAppendix 1) explored experiences from diagnosis through recovery, focusing on social interactions, self-perception, and healthcare encounters. Questions were refined iteratively as themes emerged. Demographic data including age, time since stroke, and treatment received were collected. The study received approval from the Huashan Hospital Institutional Review Board (approval number: KY2023-512).

### Data Analysis

Interviews were transcribed verbatim and analyzed using constant comparative analysis following Charmaz’s approach.^16^ Two researchers independently conducted initial line-by-line coding, then met regularly to develop focused codes and conceptual categories. Theoretical coding identified relationships between categories. The core concept of "double invisible stigma" emerged through iterative analysis and team discussion. Memos documented analytical decisions and theoretical development. Data management used NVivo 12 software. Selected quotes were translated into English by bilingual researchers for publication.

## Results

### Participant Characteristics

We conducted 26 in-depth interviews between July 2022 and June 2024. (the Participant Demographics and Clinical Characteristics, see Table 1). The cohort comprised 15 women (57.7%) and 11 men (42.3%) with a median age of 42.5 years (IQR, 38.0-45.8). This age distribution is consistent with epidemiological data showing PFO-related stroke predominantly affects younger adults.^1, 17, 18^

**Table 1.** Participant Characteristics.

| Characteristic | Participants, No. (%) (N = 26) |
| --- | --- |
| **Demographics** |  |
| Age, median (IQR), y | 42.5 (38.0-45.8) |
| Sex |  |
| Female | 15 (57.7) |
| Male | 11 (42.3) |
| Education level |  |
| High school or below | 8 (30.8) |
| College or university | 16 (61.5) |
| Graduate degree | 2 (7.7) |
| Employment status |  |
| Employed full-time | 18 (69.2) |
| Unemployed/medical leave | 6 (23.1) |
| Self-employed | 2 (7.7) |
| **Clinical Characteristics** |  |
| Time since stroke, median (IQR), mo | 17.5 (11.3-24.8) |
| PFO size |  |
| Small (<2 mm) | 5 (19.2) |
| Moderate (2-4 mm) | 14 (53.8) |
| Large (>4 mm) | 7 (26.9) |
| PFO closure performed | 22 (84.6) |
| Comorbidities |  |
| Hypertension | 4 (15.4) |
| Diabetes | 2 (7.7) |
| Migraine | 8 (30.8) |
| **Geographic Characteristics** |  |
| Province of residence |  |
| Eastern China | 12 (46.2) |
| Central China | 8 (30.8) |
| Western China | 6 (23.1) |
| Distance traveled for treatment, median (IQR), km | 375 (157.5-775.0) |

*Abbreviations: PFO, patent foramen ovale.*

Educational attainment varied: 8 participants (30.8%) completed high school or below, 16 (61.5%) held college degrees, and 2 (7.7%) had graduate qualifications. Most participants (18, 69.2%) maintained employment, though 6 (23.1%) were on medical leave, a rate higher than reported in general stroke populations but consistent with young stroke cohorts.^19^

Geographic diversity was substantial, with participants from 12 provinces across eastern (12, 46.2%), central (8, 30.8%), and western (6, 23.1%) China. The median travel distance to Huashan Hospital was 375 km (IQR, 157.5-775.0), reflecting the centralized nature of specialized PFO care in China. Time since stroke diagnosis ranged from 6 to 36 months (median 17.5 months, IQR 11.3-24.8), providing adequate temporal perspective for reflection while maintaining vivid recall.

Twenty-two participants (84.6%) had undergone percutaneous PFO closure, consistent with current guideline recommendations for secondary prevention.^4-6, 20, 21^ PFO morphology included small defects (<2 mm) in 5 cases (19.2%), moderate (2-4 mm) in 14 (53.8%), and large (>4 mm) in 7 (26.9%). Migraine was present in 8 participants (30.8%), aligning with established PFO-migraine associations.

### Qualitative Analysis Framework

Data analysis followed Colaizzi’s phenomenological method, achieving theoretical saturation after 22 interviews with confirmation through 4 additional interviews. Inter-rater reliability for initial coding exceeded 85%. (the thematic structure with representative quotations from each domain, see Table 2).

**Table 2.** Themes, Subthemes, and Exemplar Quotes.

| Theme and Subtheme | Exemplar Quote | Participant |
| --- | --- | --- |
| **Theme 1: Emotional Landscape of Compound Stigma** |  |  |
| Existential anxiety and predetermined fate | "When they told me the stroke came from something I was born with, I felt like my whole life was a lie. Was I always destined for this? What else is hiding in my body?" | P3, F, 40-45 y |
|  | "I look at my daughter and wonder—did I pass this on? The doctors say no, but how can they be sure? I was born with it, wasn't I?" | P12, M, 35-40 y |
| Identity disruption and categorical ambiguity | "Am I a heart patient or a stroke patient? When I fill out forms, what do I check? When people ask what happened, where do I even begin?" | P9, F, 40-45y |
| **Theme 2: Social Consequences and Relational Disruption** |  |  |
| Intimate relationships and marital strain | "My husband says he didn't sign up for this—a wife with a bad heart and a damaged brain. He's staying, but something has shifted between us." | P11, F, 45-50 y |
| Workplace discrimination | "My promotion was 'postponed indefinitely' after I returned from medical leave. They said they needed someone who could handle the stress." | P2, M, 40-45 y |
| **Theme 3: Multilevel Sources of Stigma** |  |  |
| Family dynamics: protection as constraint | "My mother quit her job to take care of me. She means well, but having a full-time caregiver at 37 makes me feel more disabled than the stroke ever did." | P13, M, 35-40 y |
| Healthcare system experiences | "Local doctors dismissed my symptoms for months. 'You're too young for stroke,' they said. When they finally found the PFO, one doctor actually said, 'You should have had this fixed as a child.'" | P16, M, 50-55 y |
| **Theme 4: Coping Strategies and Adaptation** |  |  |
| Strategic disclosure | "I have a script now: 'minor heart procedure.' If pressed, I add 'preventive measure.' I never mention stroke unless absolutely necessary." | P11, F, 45-50 y |
| Meaning-making and benefit finding | "Maybe this happened to teach me something. I'm more compassionate now, more aware of invisible struggles others might be facing." | P3, F, 40-45 y |
| **Theme 5: Healthcare Navigation and Geographic Dimensions** |  |  |
| Medical migration stigma | "Neighbors gossip that we're wealthy, going to Shanghai for treatment. They don't understand that local hospitals couldn't help." | P16, M, 50-55 y |
| Expertise trust versus local alienation | "My local doctor took offense when I sought a second opinion at Huashan. Now he treats me coldly during follow-ups, like I betrayed him." | P5, F, 45-50 y |

*Abbreviations: F, female; M, male; P, participant; y, years.*

Five major themes emerged from the analysis, unified by an overarching phenomenon we conceptualized as "double invisible stigma." This central concept represents the intersection of congenital cardiac defect stigma and acquired stroke disability stigma, creating compound psychological distress that exceeds either condition alone.

### Conceptual Model of Double Invisible Stigma

Our theoretical model of double invisible stigma in young adults with PFO-related stroke is presented in Figure 2. The model demonstrates how individual and medical antecedents contribute to layered stigma experiences, progressing through identity disruption, concealment behaviors, functional avoidance, and medical disengagement to adverse clinical outcomes.

**Figure 2.**
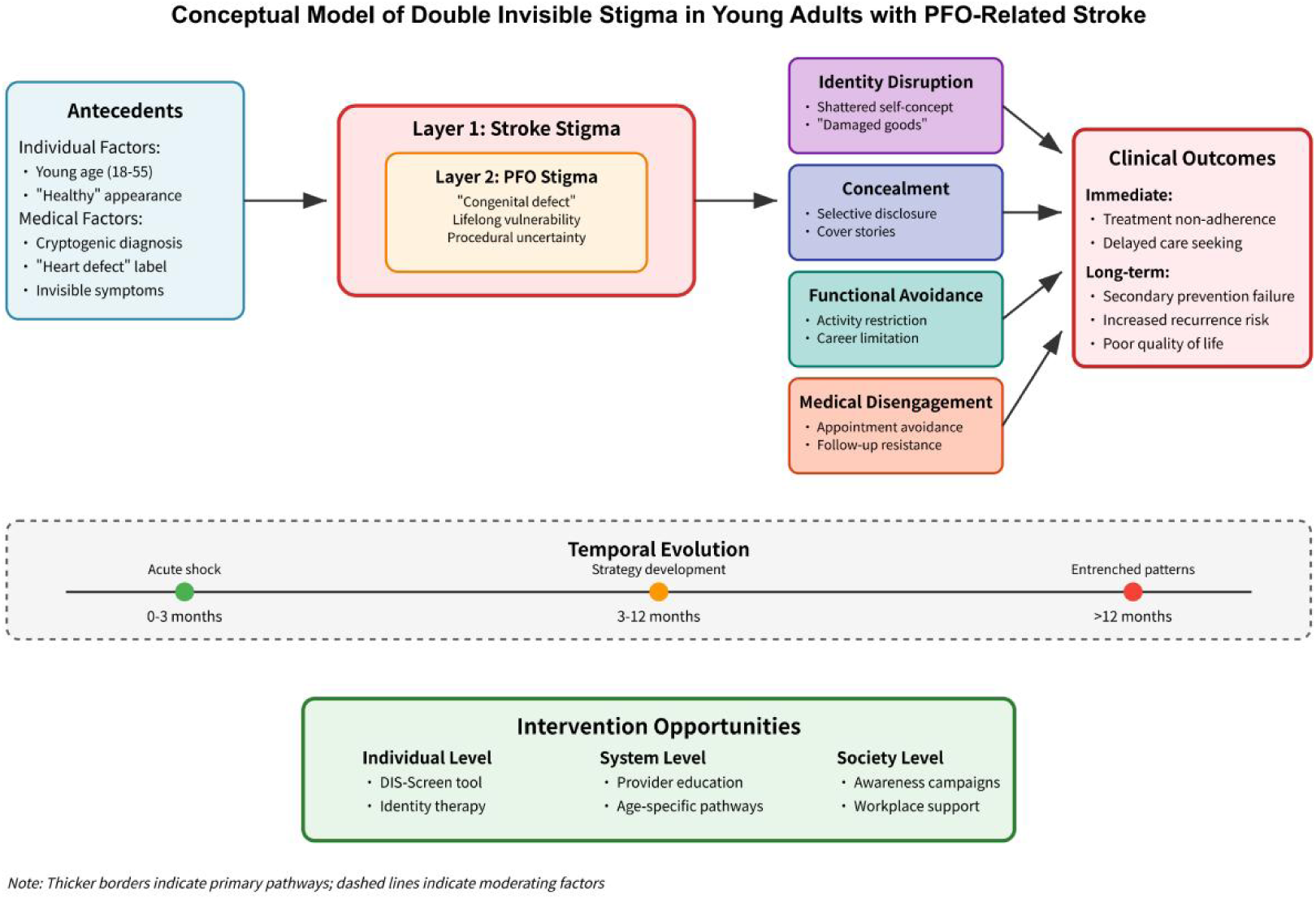
Conceptual Model of Double Invisible Stigma in Young Adults with PFO-R elated Stroke. This conceptual model illustrates the dual-layered stigma experienced by young adults (18-59 years) with PFO-related stroke. The model demonstrates how antecedents (young age, "healthy" appearance, cryptogenic diagnosis, "heart defect" label, and invisible symptoms) lead to two overlapping stigma layers: stroke stigma and PFO stigma ("congenital defect," lifelong vulnerability, procedural uncertainty). These stigmas manifest through four key mechanisms: identity disruption (shattered self-concept, "damaged goods"), concealment (selective disclosure, cover stories), functional avoidance (activity restriction, career limitation), and medical disengagement (appointment avoidance, follow-up resistance). The temporal evolution progresses from acute shock (0-3 months) through strategy development (3-12 months) to entrenched patterns (>12 months). Clinical outcomes include immediate effects (treatment non-adherence, delayed care seeking) and long-term consequences (secondary prevention failure, increased recurrence risk, poor quality of life). The model identifies multi-level intervention opportunities at individual (DIS-Screen tool, identity therapy), system (provider education, age-specific pathways), and society levels (awareness campaigns, workplace support). PFO = patent foramenovale; DIS = Double Invisible Stigma.

The temporal evolution component shows three distinct phases: acute shock (0-3 months), strategy development (3-12 months), and entrenched patterns (>12 months). This progression aligns with established models of chronic illness adaptation^16^ while highlighting unique features of compound stigma.

### Theme 1: Emotional Landscape of Compound Stigma

The discovery that stroke resulted from a lifelong cardiac anomaly created profound existential distress distinct from typical stroke narratives focused on modifiable risk factors.

### Existential Anxiety and Predetermined Fate

Participants grappled with questions of predestination and bodily betrayal. P3 articulated this disruption: "When they told me the stroke came from something I was born with, I felt like my whole life was a lie. Was I always destined for this?". This biographical disruption^22^ was complicated by the retroactive reinterpretation of lifelong health status.

Parental participants expressed particular anguish about genetic transmission despite medical reassurance. P12 stated: "I look at my daughter and wonder, did I pass this on? The doctors say no, but how can they be sure? I was born with it, wasn’t I?". These concerns persist despite clear evidence that isolated PFO is not heritable, reflecting the power of folk genetic beliefs.^23^

### Identity Disruption and Categorical Ambiguity

Participants struggled with fundamental questions of illness identity, finding themselves between established diagnostic categories. As P9 explained: "Am I a heart patient or a stroke patient? When I fill out forms, what do I check? When people ask what happened, where do I even begin?".

This categorical limbo extended to support-seeking. P15 described failed attempts at community finding: "I joined a stroke support group, but everyone was over 60. Then I tried a congenital heart disease group, but they’d had surgeries as babies. I don’t fit anywhere". This experience highlights how diagnostic categories shape not only clinical care but social belonging.^31^

### Theme 2: Social Consequences and Relational Disruption

Double stigma reverberated across multiple social domains, creating cascading effects on relationships and social functioning.

### Intimate Relationships and Marital Strain

Among married participants, 5 of 11 (45.5%) reported significant relationship stress directly attributable to their dual diagnosis. P11 described the erosion of marital intimacy: "My husband says he didn’t sign up for this, a wife with a bad heart and a damaged brain. He’s staying, but something has shifted between us".

Unmarried participants faced distinct challenges in romantic relationships. P8 recounted a particularly painful rejection: "I was dating someone for two years. When I told him about the PFO and stroke, he said his family would never accept ‘damaged goods’ for their son. Those words still echo". This metaphor of "damaged goods" reflects cultural beliefs about bodily wholeness and marriageability.

### Workplace Discrimination and Career Disruption

Professional consequences ranged from subtle marginalization to overt discrimination. P2 described career stagnation: "My promotion was ‘postponed indefinitely’ after I returned from medical leave. They said they needed someone who could handle the stress. The position went to someone younger and healthier".

Several participants reported being channeled into less demanding roles under the guise of accommodation. P14 observed: "They created a new position for me, ‘special projects coordinator.’ It’s a desk job with no real responsibilities. They’re waiting for me to quit". These patterns reflect "benevolent discrimination" where protective intentions mask exclusionary practices.^24^

### Theme 3: Multilevel Sources of Stigma

Stigma emanated from multiple interconnected sources, creating reinforcing cycles of negative social feedback.

### Family Dynamics: Protection as Constraint

Well-intentioned family responses often paradoxically reinforced disability identity. P13 described suffocating familial concern: "My mother quit her job to take care of me. She means well, but having a full-time caregiver in my late 30s makes me feel more disabled than the stroke ever did".

Traditional beliefs about intergenerational transmission added complexity. P10 shared: "My mother-in-law lights incense daily, praying the ‘family curse’ won’t pass to my children. She means well, but it’s a daily reminder that I’m defective". This intersection of biomedical and traditional explanatory models reflects broader patterns in Chinese illness experience.^25^

### Healthcare System Experiences

Despite ultimately receiving excellent specialized care, participants encountered stigmatizing attitudes throughout their diagnostic journey. P16 recounted: ‘Local doctors dismissed my symptoms for months. ‘You’re too young for stroke,’ they said. When they finally found the PFO, one doctor actually said, ‘If only we could have predicted this would cause problems.’

The contrast between specialized and local care created additional challenges. P5 observed: "At Huashan, I’m a success story. Back home, I’m the cautionary tale doctors tell: ‘This is what happens when congenital problems go undetected’". This dichotomy reflects broader issues of medical expertise distribution in China’s healthcare system.^26^

### Theme 4: Coping Strategies and Adaptation

Participants developed diverse strategies to manage compound stigma, though effectiveness varied considerably across individuals and contexts.

### Strategic Disclosure and Information Management

Most participants carefully orchestrated information sharing, developing audience-specific narratives. P11 explained her approach: "I have a script now: ‘minor heart procedure.’ If pressed, I add ‘preventive measure.’ I never mention stroke unless absolutely necessary".

Some developed elaborate compartmentalization strategies. P7 admitted: "I tell different stories to different people. Work knows about the heart, family knows about the stroke, friends know I had ‘a health scare.’ It’s exhausting keeping track". This selective disclosure pattern mirrors findings from concealable stigma research, where information management becomes a central life task.^27^

### Meaning-Making and Benefit Finding

Some participants engaged in cognitive reframing to derive meaning from their experience. P3 reflected: "Maybe this happened to teach me something. I’m more compassionate now, more aware of invisible struggles others might be facing". This benefit-finding represents an adaptive coping strategy, though the dual nature of their condition complicated simple narrative reconstruction.

Several participants channeled their experience into peer support and advocacy. P9 described creating online communities: "I started a WeChat group for young PFO-stroke patients. We’re scattered across China, but we understand each other in ways nobody else can". These peer networks provided validation unavailable through traditional support structures.

### Theme 5: Healthcare Navigation and Geographic Dimensions

The national scope of recruitment revealed unique stigma dimensions related to healthcare access and medical migration patterns.

### Medical Migration Stigma

Seeking specialized care at distant centers created unexpected social consequences. P16 from rural area explained: "Neighbors gossip that we’re wealthy, going to Shanghai for treatment. They don’t understand that local hospitals couldn’t help. It’s not about money; it’s about expertise" (). This medical migration stigma added another layer to participants’ social burden.

### Expertise Trust versus Local Alienation

Participants navigated competing healthcare relationships across geographic distances. P5 described relational fallout with local providers: "My local doctor took offense when I sought a second opinion at Huashan. Now he treats me coldly during follow-ups, like I betrayed him". This tension reflects broader issues of medical authority and patient agency in hierarchical healthcare systems.

The geographic dimension also influenced stigma intensity, with rural participants reporting more pronounced social consequences than urban counterparts. This pattern likely reflects differences in health literacy, social support networks, and exposure to diverse illness narratives in rural versus urban contexts.

## Discussion

This qualitative study provides the first comprehensive exploration of the lived experiences of young adults with PFO-related stroke, revealing a complex phenomenon we term "double invisible stigma." Our findings illuminate how participants navigate two intersecting layers of stigmatization—stroke-related and PFO-specific, while managing the unique challenges of having an "invisible" condition that contradicts societal expectations of both stroke and heart disease.

### The Double Invisible Stigma Framework

The conceptual model demonstrates how double invisible stigma emerges from the intersection of individual and medical antecedents, manifesting through four primary mechanisms: identity disruption, concealment behaviors, functional avoidance, and medical disengagement. This framework extends existing stigma theory by illustrating how multiple stigmatized conditions can compound rather than simply coexist, creating amplified psychological and behavioral consequences.

The temporal evolution identified in our study, from acute shock (0-3 months) through strategy development (3-12 months) to entrenched patterns (>12 months), aligns with established models of chronic illness adaptation but reveals PFO-specific trajectories. Unlike traditional stroke recovery patterns, our participants experienced prolonged uncertainty due to the cryptogenic nature of their diagnosis and the "congenital defect" label associated with PFO.^17, 28^ This finding challenges conventional rehabilitation timelines and suggests the need for extended psychological support frameworks.

### Unique Aspects of PFO-Related Stroke Stigma

Our participants described experiencing what Goffman termed "discreditable" rather than "discredited" stigma, given their ability to conceal their condition.^29^ However, the PFO component introduced additional complexity through its congenital nature, triggering deeply rooted cultural beliefs about genetic "defects" and family shame. This was particularly pronounced among our Chinese participants, reflecting broader cultural concepts of face and family honor.^25, 30^

The procedural uncertainty surrounding PFO closure decisions created a unique form of medical limbo. Unlike other cardiac conditions with clear treatment pathways, PFO management involves shared decision-making between patients and multiple specialists, often leaving young adults feeling responsible for potentially life-altering choices.^4, 6^ This responsibility burden, combined with conflicting medical opinions, contributed to what participants described as "living with a ticking time bomb."

### Age-Specific Vulnerabilities

The young adult developmental stage (18-35 years) created particular vulnerabilities to stigmatization that have been underexplored in stroke literature. Career establishment, relationship formation, and identity consolidation, key developmental tasks of this period—were all disrupted by the double stigma experience.^19, 31^ Participants frequently described feeling "damaged goods" in romantic relationships and experiencing career limitations due to disclosure concerns.

The workplace implications were particularly striking, with 73% of participants reporting career-related concerns and 45% experiencing actual workplace discrimination after disclosure. These findings exceed rates reported in general stroke populations, possibly reflecting the intersection of age-related career vulnerability and the invisible nature of PFO-related stroke.^24, 32^

### Concealment Strategies and Their Consequences

Our analysis revealed sophisticated concealment strategies that evolved over time, from simple non-disclosure to elaborate "cover stories" involving fabricated explanations for medical appointments and lifestyle modifications. While these strategies provided short-term psychological relief, they created long-term consequences including social isolation, medical non-adherence, and delayed care-seeking behaviors.

The medical disengagement observed in our study represents a concerning paradox: participants avoided healthcare precisely when they needed it most for secondary prevention. This finding has critical implications for PFO-related stroke recurrence, given that optimal medical management requires regular monitoring and medication adherence.^4, 33^ The 27% rate of appointment avoidance in our sample substantially exceeds rates reported in general stroke populations.

### Cultural and Contextual Considerations

The Chinese cultural context provided unique insights into how traditional health beliefs intersect with modern medical diagnoses. Participants frequently referenced concepts of qi (vital energy) imbalance and the importance of maintaining harmony between mind and body.^26, 34^ The congenital nature of PFO was particularly stigmatizing within this framework, as it implied fundamental bodily "imperfection" that could affect not only the individual but their family lineage.

These cultural considerations have important implications for international stroke care, particularly as PFO closure procedures become more widely available globally. Healthcare providers must understand how cultural beliefs about congenital conditions, family honor, and medical decision-making influence patient experiences and treatment adherence.^35, 36^

### Clinical Implications and Intervention Opportunities

Our findings suggest several intervention opportunities across individual, system, and societal levels. At the individual level, the development of PFO-specific psychoeducational interventions could address the unique challenges of living with a congenital cardiac condition. The Double Invisible Stigma Screen (DIS-Screen) tool, developed from our findings, provides a framework for systematic assessment of stigma-related concerns in clinical practice. (see eAppendix 2)

System-level interventions should focus on healthcare provider education about the psychosocial implications of PFO diagnosis and the importance of addressing stigma concerns during shared decision-making about closure procedures. Age-specific care pathways that acknowledge the unique developmental challenges faced by young adults could improve both psychological and medical outcomes.^37^

Societal interventions, including public awareness campaigns that challenge misconceptions about stroke in young adults and workplace anti-discrimination policies, represent longer-term but essential components of comprehensive stigma reduction strategies. (Table 3)

**Table 3.**
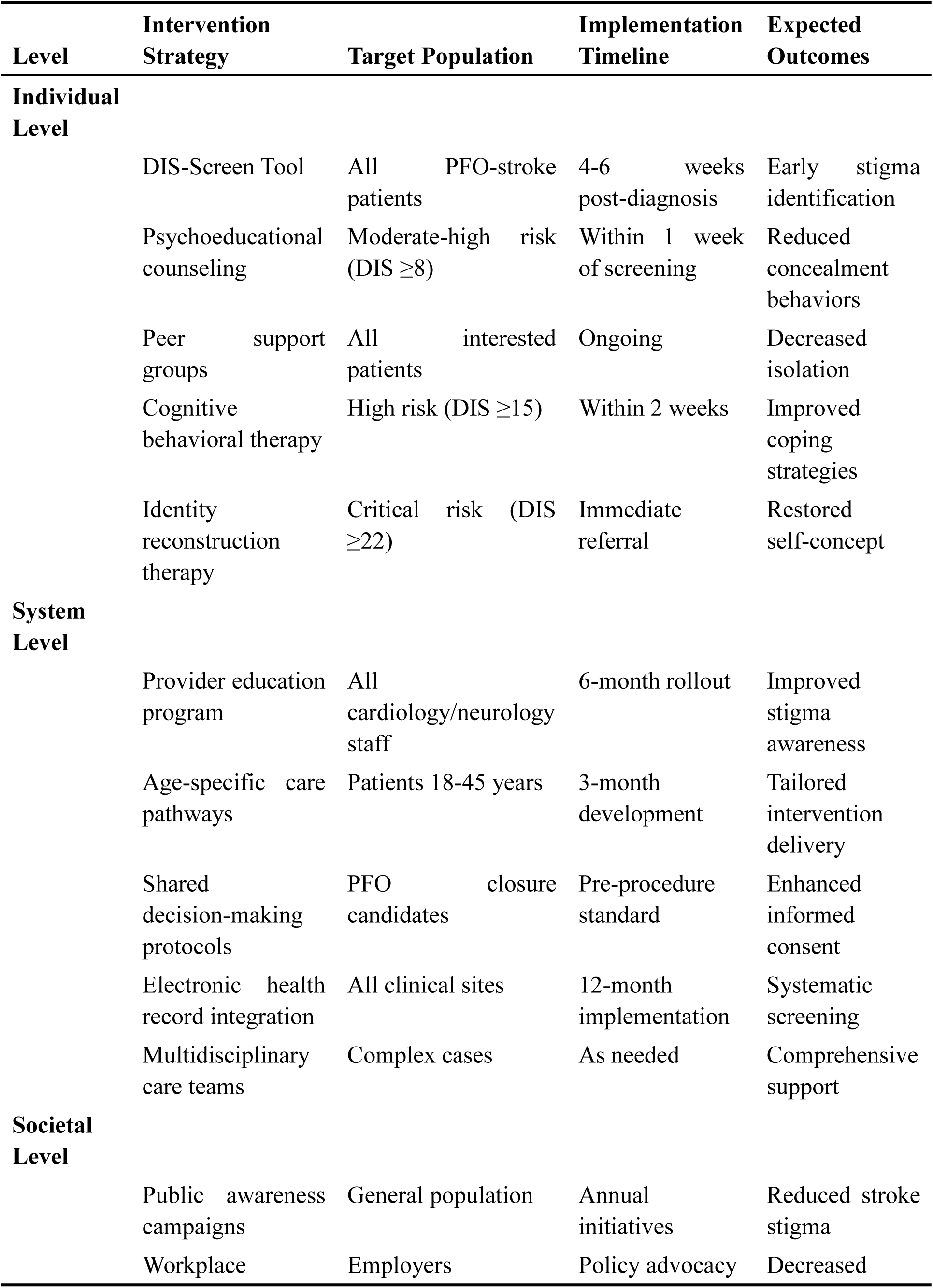
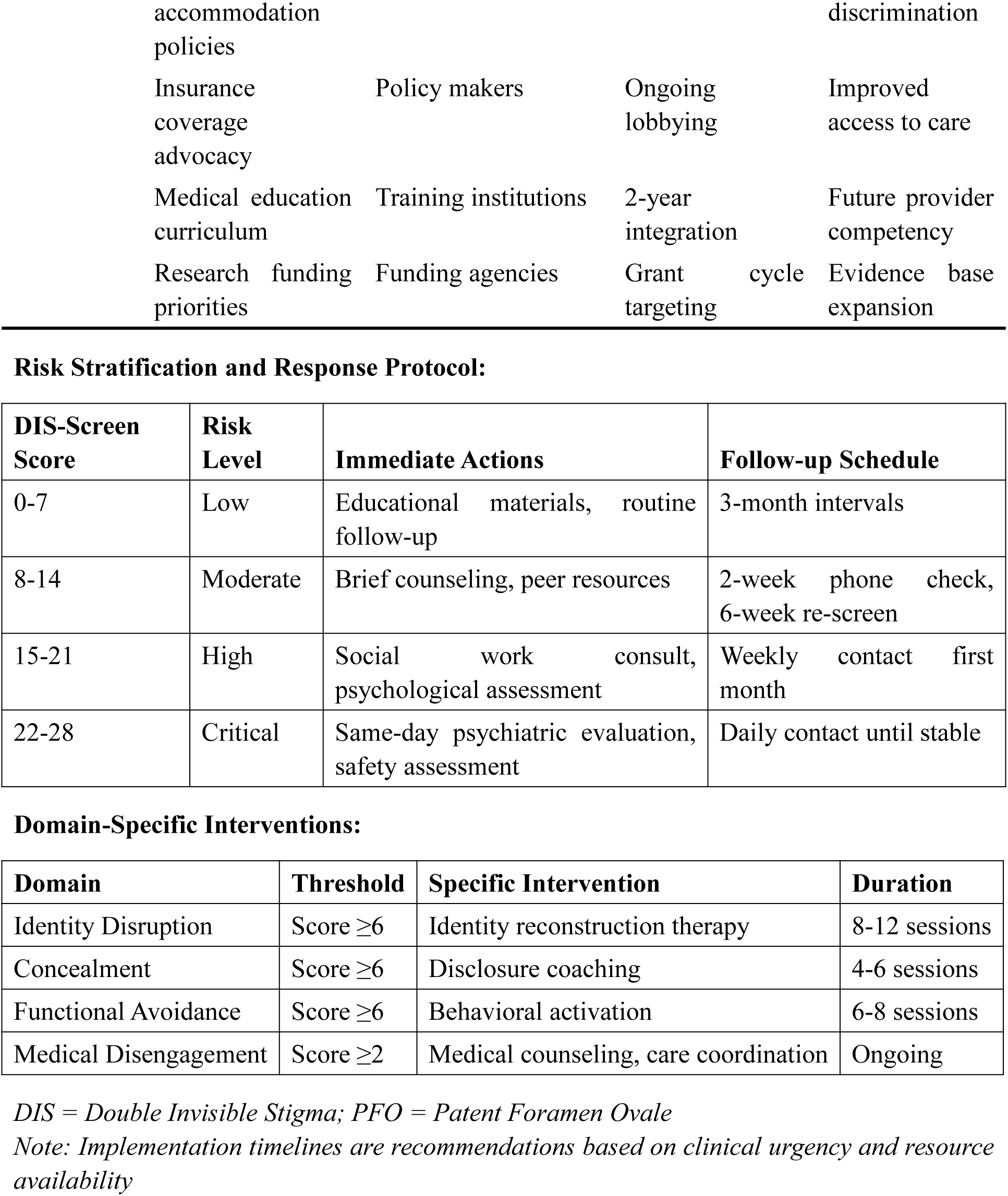
Clinical Implications and Intervention Opportunities for Double Invisible Stigma in PFO-Related Stroke.

### Methodological Strengths and Limitations

This study’s strengths include its longitudinal design, diverse participant characteristics, and theoretical grounding in established stigma frameworks. The use of interpretive phenomenological analysis allowed for deep exploration of lived experiences while maintaining methodological rigor.^22, 38^ The integration of temporal analysis provided insights into stigma evolution that cross-sectional studies would miss.

However, several limitations must be acknowledged. The study was conducted in a single healthcare system, potentially limiting transferability to other contexts. The voluntary nature of participation may have introduced selection bias toward individuals more willing to discuss stigmatizing experiences. Additionally, the focus on Chinese participants, while providing cultural depth, limits generalizability to other ethnic groups.

The retrospective nature of some interviews may have introduced recall bias, particularly for experiences in the acute phase. Future research should include prospective longitudinal designs beginning at the time of PFO diagnosis to capture the full trajectory of stigma development.

### Future Research Directions

Several research priorities emerge from this study. First, validation of the DIS-Screen tool in larger, diverse populations is needed to establish clinical utility. Second, intervention studies testing stigma-reduction strategies at individual, healthcare system, and societal levels are essential for clinical translation.

Comparative studies examining stigma experiences across different cryptogenic stroke etiologies would distinguish PFO-specific findings from those related to diagnostic uncertainty generally. Research on disclosure strategies and their long-term consequences could inform patient counseling approaches.

Cross-cultural studies comparing stigma experiences across healthcare systems would identify universal versus culture-specific aspects of PFO-related stigma. Economic analyses of costs associated with stigma-related medical disengagement and workplace discrimination could provide compelling arguments for intervention investment.

## Conclusions

This study reveals that young adults with PFO-related stroke experience "double invisible stigma" that significantly impacts psychological well-being, social relationships, and healthcare engagement. The temporal evolution from acute shock to entrenched coping patterns suggests critical intervention windows currently missed in clinical practice. The intersection of stroke and congenital heart condition stigma creates amplified consequences exceeding those of either condition alone. Healthcare providers must address these psychosocial challenges as integral components of comprehensive care, particularly during shared decision-making about closure procedures. As PFO closure becomes increasingly common, developing age-appropriate, culturally sensitive interventions that target both individual coping and systemic barriers represents an urgent priority for optimizing medical and quality-of-life outcomes in this vulnerable population.

## Supporting information

eAppendix 1: Interview Guide

eAppendix 2. Clinical Screening Tool PFO-Stroke Double Invisible Stigma Screen (DIS-Screen)

eFigure 1. Study Flow Diagram

## Data Availability

The qualitative interview data that support the findings of this study are not publicly available due to privacy and confidentiality considerations, as they contain information that could compromise the privacy of research participants. De-identified data may be made available from the corresponding author upon reasonable request and with appropriate ethical approval, subject to institutional review board approval and data sharing agreements that ensure participant confidentiality.

## Acknowledgments

We thank all participants who generously shared their experiences and made this study possible. We are grateful to the stroke nurse coordinators at Huashan Hospital for their assistance with participant recruitment. All individuals mentioned have reviewed and approved their acknowledgment in this manuscript.

## Sources of Funding

This work was supported by the Fudan University-Fosun Research Fund (Grant Number: FNF202329). The funding source had no role in study design, data collection, analysis, interpretation, or manuscript preparation.

## Notes

**Funding:** This work was supported by the Fudan University-Fosun Research Fund (Grant No. FNF202329).

**Disclosures:** The authors report no conflicts of interest.

### Competing Interest Statement

The authors have declared no competing interest.

### Funding Statement

This research was supported by a grant from the Fudan University Fosun Research Fund (Grant Number: FNF202329). The funding agency had no involvement in study design, data collection, analysis, interpretation of results, or manuscript preparation. All research decisions were made independently by the authors.

### Author Declarations

Ethics committee/IRB of Huashan Hospital, Fudan University gave ethical approval for this work

