## Supplementary material for "Double Invisible Stigma in Young Adults with PFO-Related Stroke: A Qualitative Study": eAppendix 1: Interview Guide

### Introduction and Consent

- Thank you for agreeing to participate in this study about experiences of patients with patent foramen ovale (PFO)-related stroke.
- This interview will last approximately 60-90 minutes and will be audio-recorded with your permission.
- All information will be kept confidential and anonymized in any reports or publications.
- You may skip any questions or stop the interview at any time.

### Opening Questions

1. Could you tell me a bit about yourself? (age, occupation, family situation)
2. Can you walk me through your experience of having a stroke and learning about your PFO diagnosis?

When did this happen?

What were your initial symptoms?

How was the PFO discovered?

### Illness Experience and Understanding

1. How did you understand the connection between PFO and your stroke when it was first explained to you?

What did healthcare providers tell you?

What questions did you have?

How did you explain it to family and friends?

2. How has having both a congenital heart condition and stroke affected your sense of self?

Do you see yourself differently now?

How do you describe your health condition to others?

### Stigma Experiences

1. Have you experienced any negative reactions or attitudes from others regarding your condition?

From family members?

From friends or colleagues?

From healthcare providers?

From your community?

2. Can you describe a specific situation where you felt judged or misunderstood because of your condition?

What happened?

How did it make you feel?

How did you respond?

3. Do you feel there are aspects of your condition that others don't understand or can't see?

What do you wish people understood better?

How does the "invisible" nature of your condition affect you?

### Social and Relational Impact

1. How has your condition affected your relationships?

With your spouse/partner?

With family members?

With friends?

With colleagues?

2. Has your condition affected your work or career?

Have you disclosed your condition at work?

Have you experienced any changes in responsibilities or opportunities?

What accommodations, if any, have you needed?

### Healthcare Navigation

1. Can you describe your experiences seeking treatment for your condition?

Where did you receive care?

Did you have to travel for specialized treatment?

What challenges did you face in accessing care?

2. How have healthcare providers treated you throughout this journey?

Have you felt believed and supported?

Have you encountered any dismissive attitudes?

How has your age affected how you were treated?

### Coping and Adaptation

1. How do you cope with the challenges related to your condition?

What strategies have you developed?

What or who has been most helpful?

What has been least helpful?

2. How do you decide whether and how to disclose your condition to others?

What factors influence your decision?

Can you give examples of when you chose to disclose or not?

3. Have you connected with other patients with similar conditions?

If yes, how has that been helpful?

If no, would you be interested in such connections?

### Meaning-Making and Future Outlook

1. How has this experience changed your perspective on life or health?

Has anything positive come from this experience?

What has been most challenging?

2. What are your concerns or hopes for the future?

Health-related concerns?

Life goals or plans?

### Geographic and Cultural Considerations

1. How has your location affected your experience with this condition?

Access to specialized care?

Community understanding or support?

Financial burden of seeking treatment?

2. Are there any cultural beliefs or attitudes that have influenced how you or others view your condition?

### Closing Questions

1. What advice would you give to someone newly diagnosed with PFO-related stroke?
2. Is there anything else about your experience that you would like to share that we haven't discussed?
3. What do you hope might come from research like this?

### Demographic Information (if not already covered)

- Age
- Gender
- Education level
- Occupation
- Marital status
- Province/city of residence
- Time since stroke
- Time since PFO diagnosis
- Whether PFO closure was performed
- Other health conditions
