## Supplementary material for "Double Invisible Stigma in Young Adults with PFO-Related Stroke: A Qualitative Study": eAppendix 2. Clinical Screening Tool PFO-Stroke Double Invisible Stigma Screen (DIS-Screen)

### Tool Overview

**Purpose:** Rapid identification of stigma-related psychological risk in young/middle-aged adults with PFO-related stroke
**Administration Time:** 3-5 minutes
**Setting:** Outpatient neurology/cardiology follow-up, PFO closure pre/post-procedure visits
**Administrator:** Nurse, physician, or trained medical assistant

### Screening Questions

#### Domain 1: Identity Disruption (2 items)

**1.** "Since your stroke/heart condition diagnosis, how often do you feel like you're 'damaged' or 'not normal' compared to others your age?"

□ Never (0)

□ Rarely (1)

□ Sometimes (2)

□ Often (3)

□ Always (4)

**2.** "Do you feel that having a 'hole in the heart' makes you fundamentally different from healthy people, even after treatment?"

□ Not at all (0)

□ A little (1)

□ Moderately (2)

□ Quite a bit (3)

□ Extremely (4)

#### Domain 2: Concealment Behaviors (2 items)

**3.** "In the past month, how often have you avoided telling someone about your stroke or PFO when it might have been relevant or helpful?"

□ Never (0)

□ 1-2 times (1)

□ 3-4 times (2)

□ 5-6 times (3)

□ More than 6 times (4)

**4.** "Have you ever told people you had a different health problem (like migraine or fatigue) to avoid explaining your stroke or heart condition?"

□ Never (0)

□ Once (1)

□ A few times (2)

□ Many times (3)

□ Almost always (4)

#### Domain 3: Functional Avoidance (2 items)

**5.** "Because of concerns about how others might react to your condition, have you declined or avoided work opportunities, social events, or physical activities?"

□ Not at all (0)

□ Minimally (1)

□ Somewhat (2)

□ Significantly (3)

□ Completely avoided major opportunities (4)

**6.** "How much does worry about your 'invisible' condition interfere with your sleep or daily concentration?"

□ No interference (0)

□ Mild interference (1)

□ Moderate interference (2)

□ Severe interference (3)

□ Unable to function normally (4)

#### Domain 4: Medical Engagement (1 item)

**7.** "Have you ever delayed seeking medical care or downplayed symptoms because you felt doctors might think you're 'too young' or 'look too healthy' to have real problems?"

□ Never (0)

□ Once (1)

□ 2-3 times (2)

□ 4-5 times (3)

□ More than 5 times (4)

### Scoring and Interpretation

**Total Score Range:** 0-28 points

#### Risk Stratification:

- **Low Risk (0-7 points):** Normal adjustment; routine follow-up
- **Moderate Risk (8-14 points):** Early intervention indicated
- **High Risk (15-21 points):** Comprehensive psychosocial intervention needed
- **Critical Risk (22-28 points):** Immediate mental health referral required

#### Domain-Specific Flags:

- Any single item scored ≥3: Domain-specific intervention
- Medical Engagement item ≥2: Priority medical counseling
- Total Concealment score ≥6: Disclosure coaching indicated

### Clinical Response Algorithm

#### Low Risk (0-7)

- Provide educational pamphlet on young stroke recovery
- Reinforce adaptive coping observed
- Schedule routine 3-month follow-up

#### Moderate Risk (8-14)

**Immediate Actions:**

- 15-minute stigma-focused counseling session
- Introduce peer support resources
- Provide psychoeducation on "invisible illness"

**Follow-up:**

- Phone check at 2 weeks
- Re-screen at 6 weeks
- Consider group therapy referral

#### High Risk (15-21)

**Immediate Actions:**

- Same-day social work consultation
- Formal psychological assessment within 1 week
- Begin structured intervention (CBT/ACT)
- Family meeting if consented

**Follow-up:**

- Weekly contact first month
- Bi-weekly thereafter
- Coordinate with employer/school if needed

#### Critical Risk (22-28)

**Immediate Actions:**

- Same-day psychiatric evaluation
- Safety assessment
- Intensive case management activation
- Consider partial hospitalization program

**Follow-up:**

- Daily contact until stabilized
- Comprehensive care team meeting within 48 hours

### Implementation Guide

#### When to Administer:

1. **Initial:** 4-6 weeks post-diagnosis/event
2. **Pre-procedure:** Before PFO closure
3. **Post-procedure:** 1 month after closure
4. **Routine:** Every 3-6 months first year, annually thereafter
5. **Triggered:** Any visit with mood/adherence concerns

#### How to Introduce:

"We've learned that young people with stroke and PFO often face unique emotional challenges that can affect their recovery. These brief questions help us provide better support. There are no right or wrong answers."

#### Documentation:

- Record total score and risk tier in EMR
- Flag domain-specific elevations
- Document interventions initiated
- Set automatic re-screen reminders

#### Red Flag Responses Requiring Immediate Action:

- Suicidal ideation (add PHQ-9 item 9 if suspected)
- Complete social withdrawal (item 5 = 4)
- Multiple missed appointments with concealment
- Medication non-adherence with high concealment score

### Validation Notes

*This screening tool is derived from qualitative themes and requires psychometric validation. Initial content validity established through:*

- Patient narrative analysis (n=26)
- Expert panel review (pending)
- Cognitive interviewing (pending)

*Recommended validation steps:*

1. Test-retest reliability (2-week interval)
2. Convergent validity with established stigma scales
3. Predictive validity for adherence and QOL outcomes
4. Sensitivity to change with intervention

### Quick Reference Card (for clinical use)

**DIS-Screen Quick Guide**

- 7 questions, 3-5 minutes
- Score 0-28 (higher = more stigma)
- Risk levels: Low (0-7), Moderate (8-14), High (15-21), Critical (22-28)
- Any item ≥3 = attention needed
- Re-screen: 1, 3, 6, 12 months

**Key Actions by Risk:**

- Low: Education + routine f/u
- Moderate: Brief counseling + resources
- High: Psych referral + intensive support
- Critical: Same-day psychiatric evaluation
