## Supplementary figures and images for "Double Invisible Stigma in Young Adults with PFO-Related Stroke: A Qualitative Study"

### eFigure 1. Study Flow Diagram

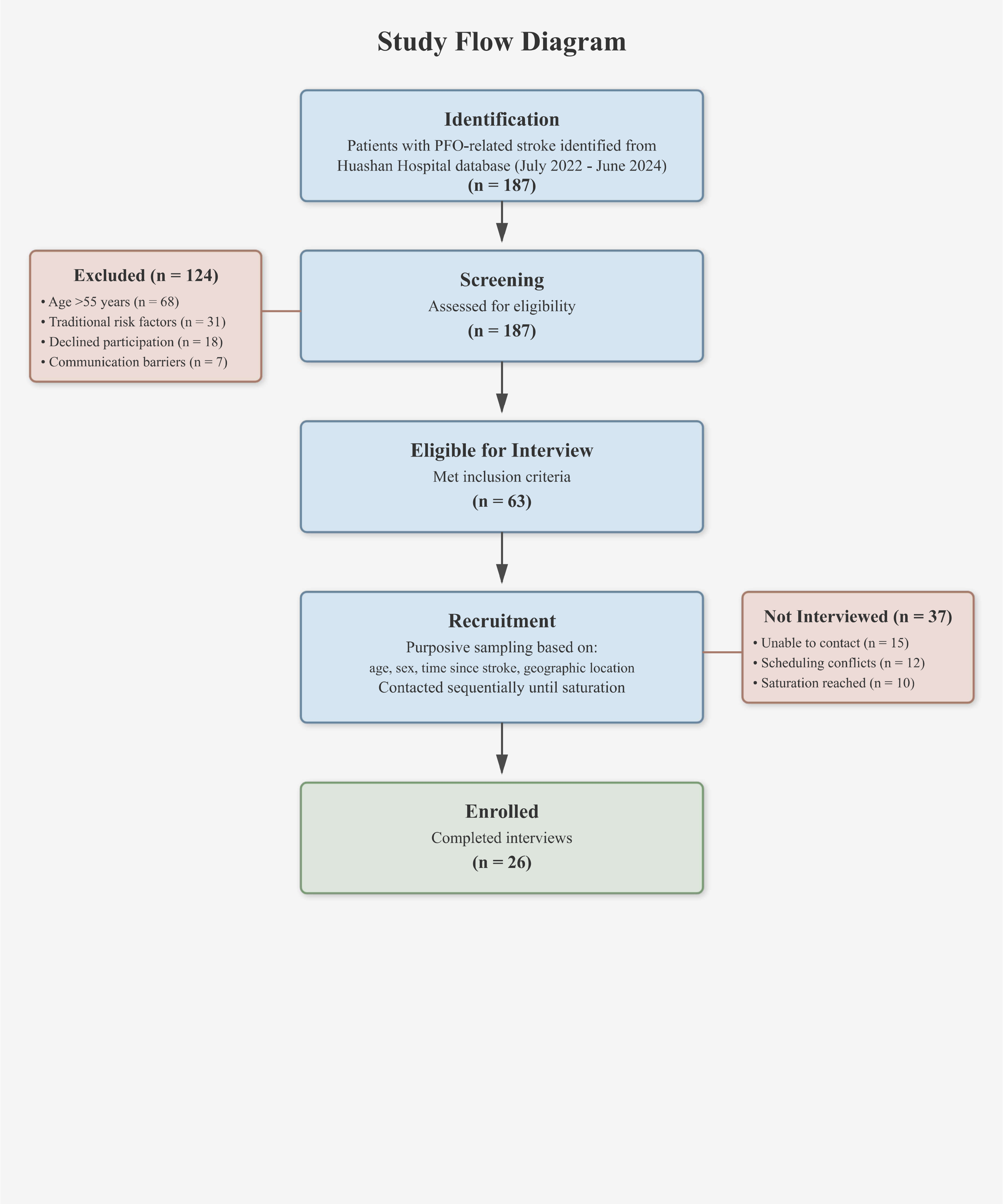
